# Validation of the Arabic version of the Orthognathic Quality of Life Questionnaire

**DOI:** 10.1101/2024.01.02.23300573

**Authors:** Shoroog Hassan Agou

## Abstract

The Orthognathic Quality of Life Questionnaire (OQLQ) was developed just over twenty years ago to measure quality of life (QoL) changes in patients with dentofacial deformities undergoing orthognathic surgery. However, despite being used in several studies conducted in Arabic-speaking nations of the Middle East and North Africa region, there has yet to be a formal psychometric evaluation of an Arabic OQLQ. The aim of this study was to develop and assess the reliability, validity, and responsiveness of an Arabic version of the OQLQ in a cohort of Saudi Arabian patients with malocclusions and dentofacial deformities. The translation of the OQLQ used an established method for adapting health questionnaires, where the questionnaire underwent translation and back-translation by bilinguals with subsequent consultation with professionals. Cronbach’s α coefficient for internal consistency was 0.92, denoting “excellent” internal consistency, and test-retest reliability was good to excellent. Overall OQOL scores were weakly but positively correlated with satisfaction with surgical treatment and were significantly lower after surgery than before surgery. The Arabic version of the OQLQ is conceptually similar to the English version, comprehensible, reliable, valid, and responsive. Future studies should now use this standardized version to ensure reliability of the results and allow comparisons between studies.

## INTRODUCTION

Orthognathic treatment describes the correction of dentofacial deformities and malocclusion with combined orthodontic and surgical management^1^. Achieving the desired clinical outcomes of harmonizing the occlusion, improving jaw function and esthetics, and ensuring long-term stability takes month to years, so the psychosocial impact of dentofacial deformities is not solely limited to the condition itself but also commitment to extensive and expensive treatment. Indeed, many patients do not even want to undergo treatment^2^. When discussing and planning the management of orthognathic treatment with patients, it is therefore necessary to have a sound evidence base of treatment efficacy based not only on clinical metrics but also in terms of how the treatment will affect the patient’s overall wellbeing, i.e., their quality of life (QoL). Recognizing the importance of the patient’s perspective in clinical practice, many QoL questionnaires have been developed for general use or for oral and maxillofacial surgery patients including the Oral Health Impact Profile (OHIP)^3^, the Short-Form Health Survey (SF-36)^4^, and many others^5^. However, most are either too general to make meaningful interpretations about the impact of orthognathic treatment or have been developed for other orofacial subsites such as pre-prosthetic surgery and dental implants^5^. Therefore, many questionnaires lack sufficient construct validity - whether the questionnaire measures what it is supposed to measure - for use in the context of orthognathic treatment.

To this end, the Orthognathic Quality of Life Questionnaire (OQLQ) was developed just over twenty years ago to measure QoL changes in patients with dentofacial deformities undergoing orthognathic surgery^6,7^, and it remains the only questionnaire for this purpose^5^. As a result, it has been used extensively in observational studies evaluating the impact of orthognathic surgery on the QoL of patients with dentofacial deformities, with a recent meta-analysis confirming the generally positive impact of the procedure^8^. The instrument has high validity and reliability^6,7^ and, given its utility in assessing patient-reported QoL outcomes after orthognathic surgery, it has been used worldwide after translation and validation in several languages including Spanish^9,10^, Serbian^11^, German^12^, Portuguese^13,14^, Chinese^15^, Dutch^16^, and Swedish^17^. Arabic versions of the OQLQ have also been used to assess QoL before or after surgery in patients in Middle East and North African (MENA) countries^18–24^. However, none of these studies performed formal psychometric evaluations of the translated versions, namely assessing the reliability (how consistent the results are), the validity (whether the questionnaire measures what it is supposed to measure), nor the responsiveness (ability to detect clinically important changes over time, even if small).

The aim of this study was to assess the reliability, validity, and responsiveness of an Arabic version of the OQLQ in a cohort of Saudi Arabian patients with malocclusions and dentofacial deformities.

## MATERIALS AND METHODS

### Translation process

The OQLQ instrument comprises 22 items and is divided in four dimensions: social aspect of dentofacial deformity (eight items); facial aesthetics (five items); oral function (five items); and awareness of dentofacial aesthetics (four items)^6,7^. The items are rated on a four-point Likert scale, and the total score ranges from 0 to 88. A higher score indicates lower quality of life^6,7^.

The translation process used an established method for adapting health questionnaires^25,26^, where the questionnaire undergoes translation and back-translation by bilinguals with subsequent consultation with professionals. The first translation into Arabic was carried out by an independent bilingual professional translator with Arabic as their first language, who was asked to maintain conceptual equivalence. Then, the first version of the Arabic translation was back-translated into English by another independent professional translator. The committee of professionals included the author, two dental interns, and both the forward and backward translators, who reviewed the translations and determined whether the translated and original versions achieved semantic, idiomatic, experiential, and conceptual equivalence. Any discrepancies were resolved, and members of the expert committee reached a consensus on all items used to generate a prefinal version of the translate questionnaire. For pilot testing, five volunteers were asked to respond to each questionnaire item and report any difficulty in interpreting or answering the questions. Finally, making any changes through piloting, the Arabic translation process was deemed valid.

### Study design, participants, and ethical approval

This study is reported according to the STROBE statement for cross-sectional studies (see S**upplementary Checklist**)^27^. This was a prospective, cross-sectional, longitudinal study of adult patients (>18 years) seeking orthognathic treatment. Other inclusion criteria were agreement to participate and mentally competent to complete a questionnaire. The Institutional Review Board of King Abdulaziz University approved the study protocol. All participants were fully informed of the study protocol and provided written informed consent.

Recruitment was carried out between January 2021 and December 2022 in Jeddah, Saudi Arabia. Participants in the longitudinal sample (n=74) attended a private orthodontic practice for orthognathic treatment. Each patient was invited to complete a questionnaire (**Supplementary Questionnaire**) that included basic information about the reasons for surgery, the OQLQ itself, four questions asking to what extent pain/discomfort, chewing, appearance, and speaking had been affected by the surgery, and a visual-analogue scale (VAS) to rate the overall satisfaction with orthognathic treatment. Patients were asked to complete the questionnaire at various times throughout the treatment journey: before starting treatment, after orthodontic treatment but before surgery, after surgery (at follow-up), on debonding, and after one year with retainers. A subgroup of patients (n=12) completed the questionnaire both before surgery (before treatment or after orthodontic treatment) and after surgery (after one year with retainers). To examine test-retest reliability, 15 patients attending King Abdulaziz University Hospital for orthognathic treatment also completed the same questionnaire twice with a two-week interval.

### Reliability and validity assessment and statistical analysis

Data were analyzed using SPSS v22 (IBM Statistics, Chicago, IL). Normality of continuous data was assessed with the Shapiro Wilk test. Means (standard deviation, SD) and medians (interquartile range, IQR) are presented for quantitative data, while frequencies and percentages are presented for all qualitative data. The chi-square test was used to compare proportions of two variables. The Mann-Whitney U test was used to compare QoL scores between two groups and the Wilcoxon signed-rank test was used to compare QoL scores between three or more groups. Internal consistency was assessed in the entire study population using Cronbach’s α coefficient, which are considered good if ≥ 0.70 (excellent if ≥ 0.90)^28^. Test-retest reliability was determined with Spearman’s rank correlations^28^, as were correlations between satisfaction levels and QoL. A p-value <0.05 was considered statistically significant.

## RESULTS

### Baseline characteristics of the study population

The baseline characteristics of the study population are shown in **Table 1**. Of the 74 participants, 46 (62.2%) were female, and the average age was 30 ± 4.6 years (range 20 – 44). Most patients were seeking treatment due to concerns with both orofacial appearance and function, which necessitated a range of surgical solutions to one or both jaws (83.8% of cases) that most commonly included asymmetric mandibular setback (51.4%) and maxillary advancement (25.7%).

**Table 1.** Baseline characteristics of the study population (cross-sectional data, n=74).

| Variable |  | Frequency | Percent |
| --- | --- | --- | --- |
| Sex | Male | 28 | 37.8 |
|  | Female | 46 | 62.2 |
| Age (Years) | Mean ± SD (range) | $30 \pm 4.6$ (20-44) | |
| Angle's classification of deformity | Class I | 5 | 6.8 |
|  | Class II | 13 | 17.6 |
|  | Class III | 47 | 63.5 |
| Facial type | Mesocephalic | 24 | 32.4 |
|  | Brachycephalic | 4 | 5.4 |
|  | Dolichocephalic | 25 | 33.8 |
| Vertical | Deep bite | 11 | 14.9 |
|  | Anterior overbite | 17 | 23 |
|  | Posterior cross bite | 3 | 4.1 |
|  | Vertical maxillary excess | 5 | 6.8 |
|  | Multiple responses | 12 | 16.2 |
| Transverse | Unilateral | 5 | 6.8 |
|  | Bilateral posterior cross bite | 18 | 24.3 |
| Surgical treatment details | Maxillary advancement | 19 | 25.7 |
|  | Maxillary segmental setback | 1 | 1.4 |
|  | Maxillary impaction | 16 | 21.6 |
|  | Advancement and impaction | 18 | 24.3 |
|  | Multiple responses | 8 | 5.4 |
|  | Mandibular advancement | 20 | 27 |
|  | Mandibular segmental setback | 5 | 6.8 |
|  | Mandibular asymmetric setback | 38 | 51.4 |
| Jaws operated on | Maxilla | 1 | 1.4 |
|  | Mandible | 2 | 2.7 |
|  | Both | 62 | 83.8 |
| Any other plastic surgical procedures performed? | Yes | 37 | 50 |
|  | No | 17 | 23 |
| Reasons for surgery | Appearance | 15 | 20.3 |
|  | Function | 17 | 23 |
|  | Confidence | 3 | 4.1 |
|  | Appearance and function | 16 | 21.6 |
|  | Appearance and confidence | 1 | 1.4 |
|  | Function and confidence | 2 | 2.7 |
|  | All | 17 | 23 |

### Reliability of the Arabic version of the OQLQ

Seventy-four patients completed the Arabic version of the OQLQ either before or after surgery. Across the entire study population (n=74), Cronbach’s α coefficient for internal consistency was 0.92, denoting “excellent” internal consistency.

Fifteen individuals from a separate cohort of patients attending the University Hospital were invited to complete the questionnaire and repeat it two weeks later. Test-retest reliability assessed in these 15 individuals was between 0.601 and 0.879 (**Table 2**). All questions showed “excellent” test-retest reliability (Spearman’s correlation ≥ 0.75^28^) apart from for the awareness of dentofacial deformity, which was “good” (Spearman’s correlation = 0.601).

**Table 2.** Test-retest reliability of the OQLQ (n=15, Spearman’s rank correlation coefficient).

| Domain | Correlation coefficient | Significance |
| --- | --- | --- |
| Overall | 0.859 | <0.001 |
| Social aspects of deformity | 0.849 | <0.001 |
| Facial esthetics | 0.849 | <0.001 |
| Oral function | 0.879 | <0.001 |
| Awareness of dentofacial deformity | 0.601 | 0.018 |

### Validity of the Arabic version of the OQLQ

Validity of the OQLQ was measured with respect to two variables that might be expected to be related to QoL in patients with dentofacial deformity undergoing orthognathic surgery: subjective effect on appearance after surgery (measured using a Likert scale) and overall satisfaction with surgery (measured with a VAS). The overall OQOL and facial esthetics domain scores were weakly but positively correlated with VAS-recorded satisfaction with surgical treatment (**Table 3**). Further supporting the hypothesis that OQLQ is valid for measuring QoL in orthognathic surgery patients, overall OQLQ scores were significantly lower (better QoL) in patients who felt that their overall appearance was better after surgery (median 9, IQR 7-11) compared with those felt that their appearance was only a little better (median 18.5, IQR 13.5-24) or a lot worse (median 15, IQR 5-35) after surgery (p < 0.001, Wilcoxon signed-rank test).

**Table 3.** Correlation between VAS responses for satisfaction with surgical treatment and OQOL scores (n = 74, Spearman’s rank correlation coefficient).

| Domain | Correlation coefficient |
| --- | --- |
| Overall | 0.34* |
| Social aspects of deformity | 0.35 |
| Facial esthetics | 0.437* |
| Oral function | 0.14 |
| Awareness of dentofacial deformity | -0.05 |

\*Correlation significant at the 0.05 level

### Responsiveness of the Arabic version of the OQLQ

The responsiveness of the Arabic version of the OQLQ was examined by comparing scores before and after surgery in both the entire cohort (n=74, completed questionnaire either before or after surgery) and in a subset of patients who completed the questionnaire both before and after surgery (n=12) (**Table 4**). For both groups, overall OQLQ scores were significantly lower after surgery than before surgery (p < 0.0001 and p = 0.09, respectively). In addition, in the paired analysis, scores for the facial esthetics domain were significantly lower after surgery (p=0.03).

**Table 4.** Responsiveness of the Arabic version of the OQLQ.

|  | All data |  |  | Paired data |  |  |
| --- | --- | --- | --- | --- | --- | --- |
|  | Before surgery, median (IQR) (n=74) | After surgery, median (IQR) (n=74) | p-value | Before surgery, median (IQR) (n=12) | After surgery, median (IQR) (n=12) | p-value |
| Overall | 34 (21 - 42) | 10 (4 - 28) | **<0.0001** | 22 (14.5 - 50.5) | 11.5 (9.5 - 21) | **0.09** |
| Social aspects of deformity | 5 (3 - 9) | 3 (1 - 7.5) | 0.279 | 5.5 (0 - 13) | 1.5 (0 - 6) | 0.236 |
| Facial esthetics | 8 (5 - 15) | 5 (2 - 9) | 0.159 | 9.5 (6 - 14.5) | 6 (2.5 - 8.5) | **0.03** |
| Oral function | 6 (2 - 11) | 4 (2 - 7) | 0.343 | 4.5 (3 - 10.5) | 2.5 (0.5 - 7.5) | 0.327 |
| Awareness of dentofacial deformity | 6 (2 - 11) | 6 (2 - 11) | 0.952 | 6 (1.5 - 9) | 3 (0.5 - 6) | 0.265 |

## DISCUSSION

This study developed and formally validated an Arabic version of the OQLQ. The data – collected from a relatively large sample of consecutive patients attending a private practice for orthognathic treatment - suggest that the translated version is conceptually similar to the English version, comprehensible, reliable, valid, and responsive. The cohort represented a range of clinical presentations and motivations for undergoing orthognathic treatment and, confirming the results of previous studies, showed an overall positive effect on QoL from treatment, with a mean difference of 24 points, similar to the mean difference of 20 OQLQ points resulting from treatment reported in a previous meta-analysis^8^.

As seen with the original English version and other translations^6,7,9–11,13–17^, the Arabic OQLQ showed excellent reliability, both in terms of internal consistency (Cronbach α > 0.9) and test-retest reliability, which was also excellent (Spearman’s rank correlation >0.75) overall and for three of the four domains. Only “awareness of dentofacial deformity” had a slightly lower (but still good) reliability, most likely due to the relatively small number (n=15) of pretreatment patients given the questionnaire twice.

The construct validity was tested using two variables related to QoL in patients with dentofacial deformity undergoing orthognathic surgery: subjective effect on appearance after surgery and overall satisfaction with surgery. While the exact relationship between global measures of satisfaction with surgery and the instrument are difficult to predict, the constructs are sufficiently related to expect a correlation; indeed, this was confirmed. Both metrics were associated with the overall OQLQ, suggesting adequate construct validity of the Arabic OQLQ. The facial esthetics subdomain was positively and significantly correlated with satisfaction with surgical treatment, which might be expected, since facial esthetics has a profound impact on quality of life, particularly self-esteem^29^. However, correlations between the “social aspects of deformity”, “oral function”, and “awareness of dentofacial deformity” subdomains and satisfaction with surgical treatment were not significant, mirroring the results of the original validity testing, which showed the lowest correlations between VAS scores and the “oral function”, and “awareness of dentofacial deformity” subdomains^7^.

Previous translations and validations of the OQLQ have failed to measure responsiveness – the ability to detect clinically important changes over time – which is the most important metric to measure if a patient-reported outcome instrument is to have clinical utility. Here, responsiveness was tested through group comparisons of patients completing the questionnaire either before or after surgery and in a subset of patients who completed the questionnaire both before and after surgery (n=12). For both groups, overall OQLQ scores were significantly lower after surgery than before surgery, similar in magnitude to OQLQ decreases observed in meta-analysis (24 vs 20 points)^8^. For the entire cohort, there were no significant decreases in OQLQ scores for any of the four subdomains, which may be due to the grouping of questionnaire responses at two timepoints before surgery (on assessment and after orthodontic treatment) and three timepoints after surgery (after surgery (at follow-up), on debonding, and one year with retainers). In the paired analysis, mirroring the validity analysis, the facial esthetics subdomain was significantly improved after surgery; given that patients are likely to focus on esthetics after undergoing a procedure to alter a facial deformity, this is perhaps unsurprising. Indeed, the responsiveness testing of the original questionnaire similarly found that not all subdomains showed favorable changes after surgery^7^. The current results may also have been limited by the small number of patients in the paired analysis. Regardless, the Arabic OQLQ sensitively measures changes in QoL related to orthognathic treatment and can therefore be considered fit for purpose.

This validated version of the OQLQ is expected to improve future QoL studies in Arabic-speaking nations. Our responsiveness testing adds to a body of evidence from the MENA region showing that orthognathic treatment improves QoL. Previous studies from Saudi Arabia^18^, Kuwait^19^, Morocco^20^, Egypt^24^, and Jordan^21^ all found that OQLQ total and subdomain scores improved after orthognathic surgery. Other comparisons using the OQLQ in the MENA region included a report of higher OQLQ scores in Jordanian patients with post-surgical temporomandibular disorders^23^. There has also been a cross-cultural comparison of OQLQ scores between Jordanian and British patients^22^, which revealed a significant difference for oral function (poorer QoL in the Jordanian cohort). Although the authors suggested that this difference could be due to culture or systemic healthcare differences, it could also have been due to understanding of the questionnaire, underscoring the value of the current study.

This study has several limitations. As noted above, the test-retest validation cohort and the pre- and post-surgery paired cohort were both small, so the analyses may have been underpowered. While the comparison of OQLQ results with surgical satisfaction suggested good construct validity, convergent and discriminant validity would ideally be tested through comparison with other validated instruments such as the OHIP14^30^, Jaw Functional Limitation Scale (JFLS)^31^, or Orofacial Esthetic Scale (OES)^32^. This study did not directly test comprehension of questionnaire, instead inferring cultural appropriateness and understandability through analysis of the reliability and clinically-correlated responsiveness data, which suggest that this Arabic questionnaire is fit for purpose. Finally, the sample was taken from one private practice in Saudi Arabia and therefore may not be representative of the population as a whole.

Nevertheless, despite twenty years of use, a formally validated version of the OQLQ has hitherto been lacking. This study fills this gap and provides a psychometrically tested Arabic version of the OQLQ that is conceptually similar to the English version, comprehensible, reliable, valid, and responsive. Future studies should now use this standardized version to ensure reliability of the results and allow comparisons between studies.

## Data Availability

All data produced in the present work are contained in the manuscript

## Notes

### Competing Interest Statement

The authors have declared no competing interest.

### Funding Statement

This study did not receive any funding

### Author Declarations

Ethics committee/IRB of the Faculty of Dentistry at King Abdulaziz University gave ethical approval for this work

